# Forecasting Covid-19 Infections and Deaths Horizon in Egypt

**DOI:** 10.1101/2020.09.28.20202911

**Authors:** Shereen Nosier, Reham Salah

## Abstract

The coronavirus Covid-19 pandemic is defining a global health crisis, which is the hugest challenge the world has faced since World War II. Accordingly, the global economy as well is facing the worst economic catastrophe since the 1930s Great Depression. The case in Egypt is similar to the rest of the world. Despite being threatened by GDP decline and income losses; the Egyptian government has reacted early to restrain the pandemic outbreak. By mid-March, many measures had been undertaken to contain the spread of the virus. More than three months after imposing them, Egypt began lifting many of the restrictions put in place to curb the spread of coronavirus. Predictions of the potential spread of Covid-19 based on time series Auto Regressive Integrated Moving Average (ARIMA) and econometric Autoregressive-Distributed Lag (ARDL) forecasting models are utilized in this paper for designing and/or evaluating countermeasures. The aim of this study is threefold, first using the most recent available data to find the best prediction models for daily cases and death in Egypt and forecast them up to 7 November 2020. Second, to analyze the effect of mobility on the incidence of the pandemic using Google Community Mobility Reports (GCMR) to evaluate the results of easing lockdown restrictions. Finally, providing some recommendations that may help lessen the spread of the virus and eradicate new deaths as possible. The results revealed that mobility of population is affecting the incidence of new cases of Covid-19 significantly over the period of the study. Additionally, the total number of infections on November 7 2020 is expected to reach 102,352 cases, while the total death toll is predicted to be 5,938 according to the most accurate methods of forecasting. Accordingly, in order to sustain the predicted flat pandemic curve, many restrictions must be continued and emergency mechanisms need to be considered. For instance, adhering to the precautions of social distancing advised by the health minister and the declared hygiene rules to ensure that infection is prevented or transmitted is necessary. Besides, being prepared with re-imposing lockdown strategies and health system support are essential among others. It should also be noted that this expected pattern can shift, yet that depends on people’s actions.

## 1. Introduction

Since the mid of December 2019, the novel coronavirus (Severe Acute Respiratory Syndrome 2 “SARS-CoV-2”) from Wuhan, China, has continued to spread across the globe and turned into a pandemic currently known as Covid-19 (Yousaf, et al., 2020; Kumar, et al., 2020). Because of the rapid increase of the number of infections worldwide and the absence of vaccines, the contagious disease has turned the world upside down. In fact, as of 15 August 2020, Coronavirus has infected 21.63 million persons, and killed more than 769 thousand people with 3.56% death to infection rate. On the other hand, 14.34 million have been recovered with 66.31% recovery rate as of the same date. A month later on 16 September, the virus has infected 29.73 million individuals and the death toll reached 939,289 resulting in a mortality rate of 3.16%. Additionally, on the same date the number of recovered people has reached 21.54 million, so that the recovery rate increased to 72.46%. That is to say, the virus may have become less lethal globally as the death rate has decreased by 0.5% and the recovery rate has risen by 6.15%. To add more, due to the Covid-19 pandemic, everything has been impacted around the world. In order to prevent further transmission, strong preventive measures had been taken such as implementing lockdown and international travel bans. Despite being under lockdown for a long time of uncertainty, things are slowly easing back leading to a decision of reopening economies and relaxing movement restrictions to avoid poverty increase and job losses.

Indeed, the global economy is facing the worst economic catastrophe since the 1930s Great Depression (Breisinger, et al., 2020). The case in Egypt is not different from the rest of the world. Despite being threatened by GDP decline and income losses, the Egyptian government has reacted early to restrain the pandemic outbreak. By mid-March, many measures had been undertaken to contain the spread of the virus, including travel bans, a nighttime curfew, social gatherings ban, and the closure of some organizations, religious institutions and schools. More than three months after imposing them, Egypt began lifting many of the restrictions put in place to curb the spread of coronavirus.

Due to the fact that decisions made now and in the months to come will be some of the most important in addition to the fears of a possible powerful second wave, it is urgent that the government making those decisions has access to the best information available regarding the possible trajectory of the pandemic. Therefore, predictions on the potential spread of Covid-19 based on econometrics models can be important tools for designing and/or evaluating countermeasures. In the last few months, researchers have developed or utilized existing statistical methods to predict the number of cases and deaths (Yousaf, et al., 2020). Additionally, the released Google Community Mobility Reports (GCMR) offers a novel opportunity to investigate the relationship between mobility patterns and spread or transmission of Covid-19 (Paez, 2020). Consequently, the aim of this study is threefold, first using the most recent available data to find the best prediction models for cases and death in Egypt and forecast them. Due to the fact of the dynamic nature of the pandemic, more and continuous epidemiological models are needed for forecasting. Second, to analyze the impact of mobility on the incidence of the pandemic using GCMR to evaluate the results of easing lockdown restrictions. Finally, providing some recommendations that may help lessen the spread of the virus and eradicate new deaths as possible.

## 2. Literature Review

Despite the importance of forecasting the possible trajectory of the pandemic, as far as we know, the studies conducting predictions of cases and deaths in Egypt are still parsimonious and not timely updated to reflect the most recent situation. Saba and Elsheikh (2020) forecasted the possible number of cases in Egypt using statistical and artificial intelligence methods namely; Auto Regressive Integrated Moving Average (ARIMA) and Nonlinear Autoregressive Artificial Neural Networks. The study utilized the reported cases from 1 March to 9 May 2020 to forecast one month ahead until 8 June 2020. An increase of 280% in cases was estimated during May 2020. Similarly, El-Ghitany (2020) conducted a short-term prediction for the epidemic situation in Egypt based on data from February 14 until April 18, 2020. The researcher applied the exponential growth rate model to forecast the daily cases from 19 April to 6 June. The results implicated that by late May, infections are expected to reach more than 20,000 then starts to decline. In their study, Elmousalami and Hassanien (2020) provided daily forecasting models of Covid-19 cases by means of different time series models such as moving average (MA), weighted moving average, and single exponential smoothing. The results forecasted that the number of confirmed cases in Egypt would be doubled up to 4-folds in April. That is, they suggested that the number of coronavirus cases grows exponentially in Egypt.

Additionally, Anwar and AbdelHafez (2020) foreseen the expectant timing of coronavirus peak in Egypt. They employed the epidemic online calculator tool, which apply the Susceptible, Exposed, Infective and Removed model. In their study, the daily reports for the period 14 February to 11 May 2020 was utilized. The results indicated the number of hospitalized cases is predicted to reach a peak of 20,126 in the middle of June. While, a total of 12,303 deaths was expected. The research also claimed that the quarantine restrictions should be kept until the end of June 2020. Likewise, Tachy Health team applied a three-compartmental Susceptible-Infected-Recovered/Removed time-dependent epidemiological model, to examine the epidemic curve in four Middle East and North Africa (MENA) countries namely; Egypt, UAE, Saudi Arabia, and Algeria. For Egypt, the results anticipated a probable peak 6 June 2020 assuming that 5% of the population is susceptible, reproduction number of 2.6 and a 1/14 recovery rate. Another two scenarios were to reach the highest number of infections at 18 June or 16 July based on different assumptions. However, no studies was found that made use of GCMR in Egypt.

## 3. Descriptive Analysis

This section provides real-time actual data about the confirmed cases and death of Covid-19 in Egypt since the outbreak of the virus on mid-February 2020 until mid-September 2020. In addition, a track of Egyptians mobility and infections is presented.

### 3.1 Cases and Deaths Trends

As depicted by Figure 1, on August 15, 2020, the total number of confirmed infections nationwide in Egypt reached 96,336 since the detection of the first one on 14 February. One month later, the number of infections has risen to 101,009. Further, the death toll has been raised to 5,141 and 5,648 deaths on 15 August and 16 September 2020 respectively. On the other hand, the recovered cases has increased to 58,835 individuals on August 15 and has reached 85,745 on 16 September. Therefore, the infection rate was 0.09% (Population =102,334,404) until mid-August, the recovery rate was 61.07% and the mortality rate was 5.34%. At that time, in comparison to the global values, the recovery rate in Egypt was lower than its global counterpart by 5.24%. While, the mortality rate was 1.78% higher in Egypt. During the last month, the infection rate in Egypt has increased by 0.29% whilst the death rate has fallen by 0.27% and the recovery rate has increased by 23.54%.

**Figure 1:**
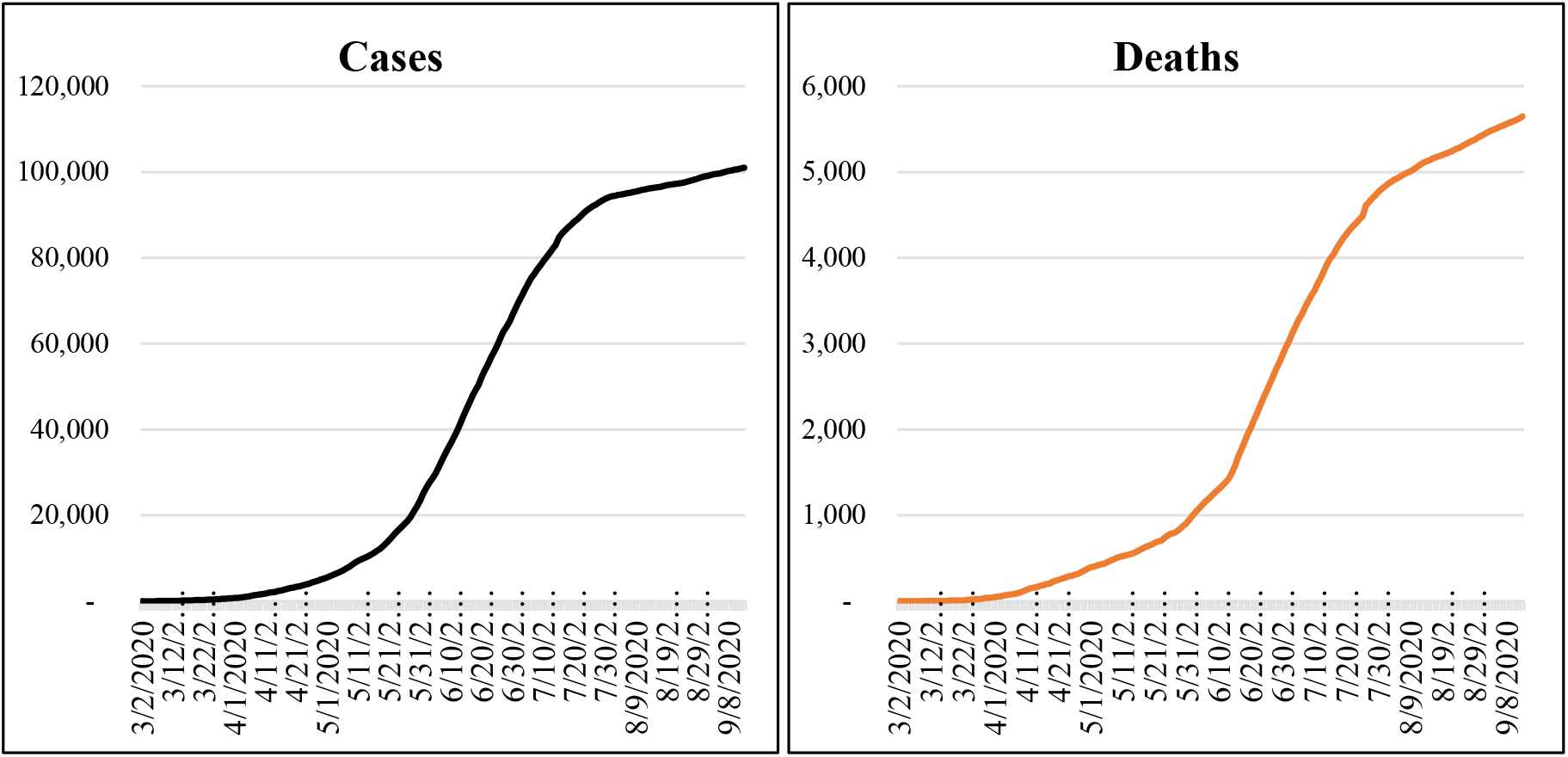
Total Confirmed Cases and Death of Covid-19

Moreover, the daily cases started to fall since July 24, 2020. In addition, the highest numbers of infections were 1,774 on 20 June 2020. Speaking of the number of death, it reached a maximum of 97 deaths on June 16, 2020. It is worth mentioning that, the number of deaths has been decreasing from 18 July 2020.

### 3.2 Mobility Trends

Obviously Egypt has experienced a long list of closures and policies aimed at encouraging citizens to stay at home. Consequently, the levels of mobility changed starting from 12 March 2020 when the government started imposing some restrictions such as schools and universities closing. At that time, the reported cases were 7 solely. Following, a night-time curfew has been imposed since March 24, 2020. Additionally, the Ministry of Awqaf [Endowment] declared the closure of the mosques and the places of worship nationwide until further notice. Figure 3 depicts how mobility to diferent destinations has altered in terms of percentage change from baseline.

**Figure 2:**
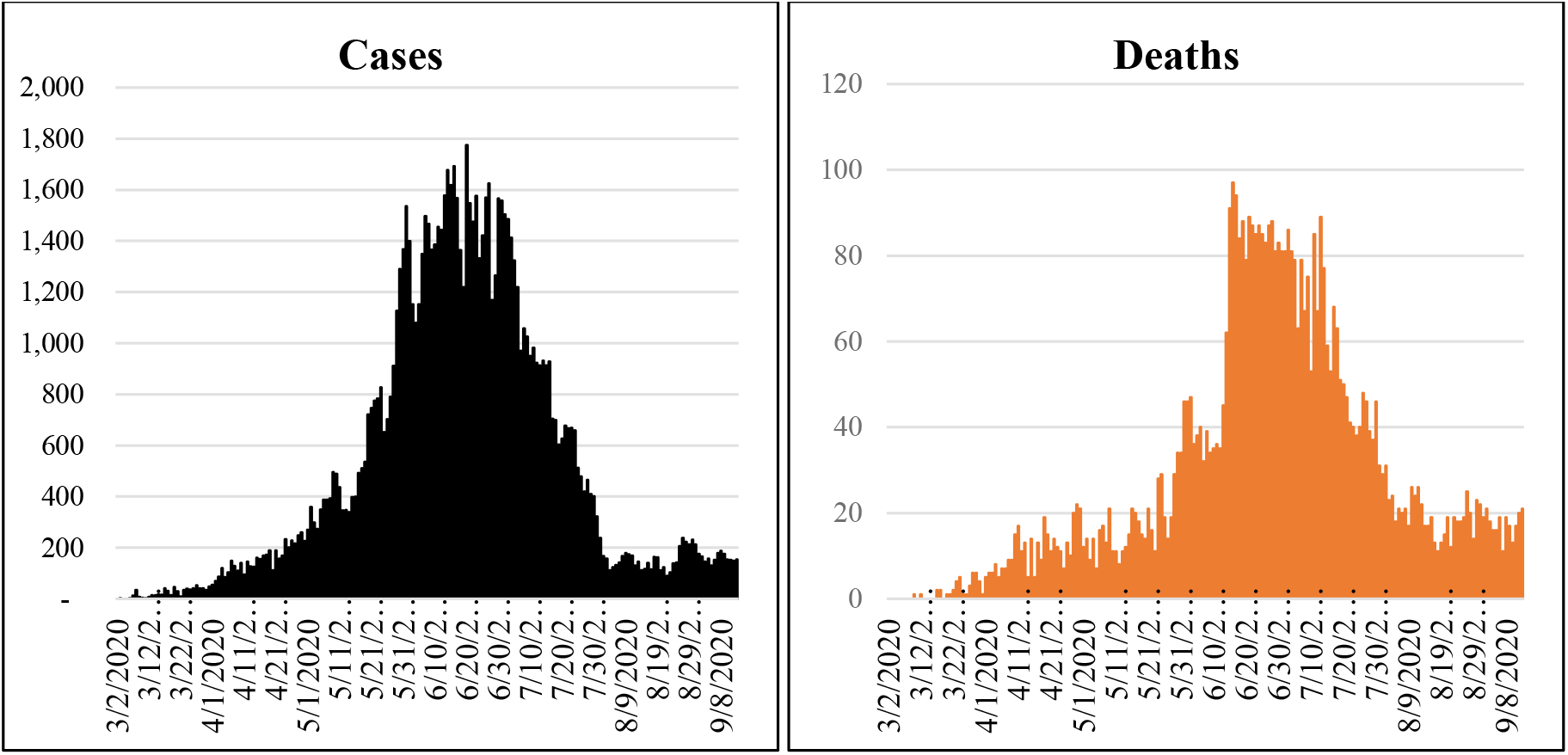
Daily Confirmed Cases and Death of Covid-19

**Figure 3:**
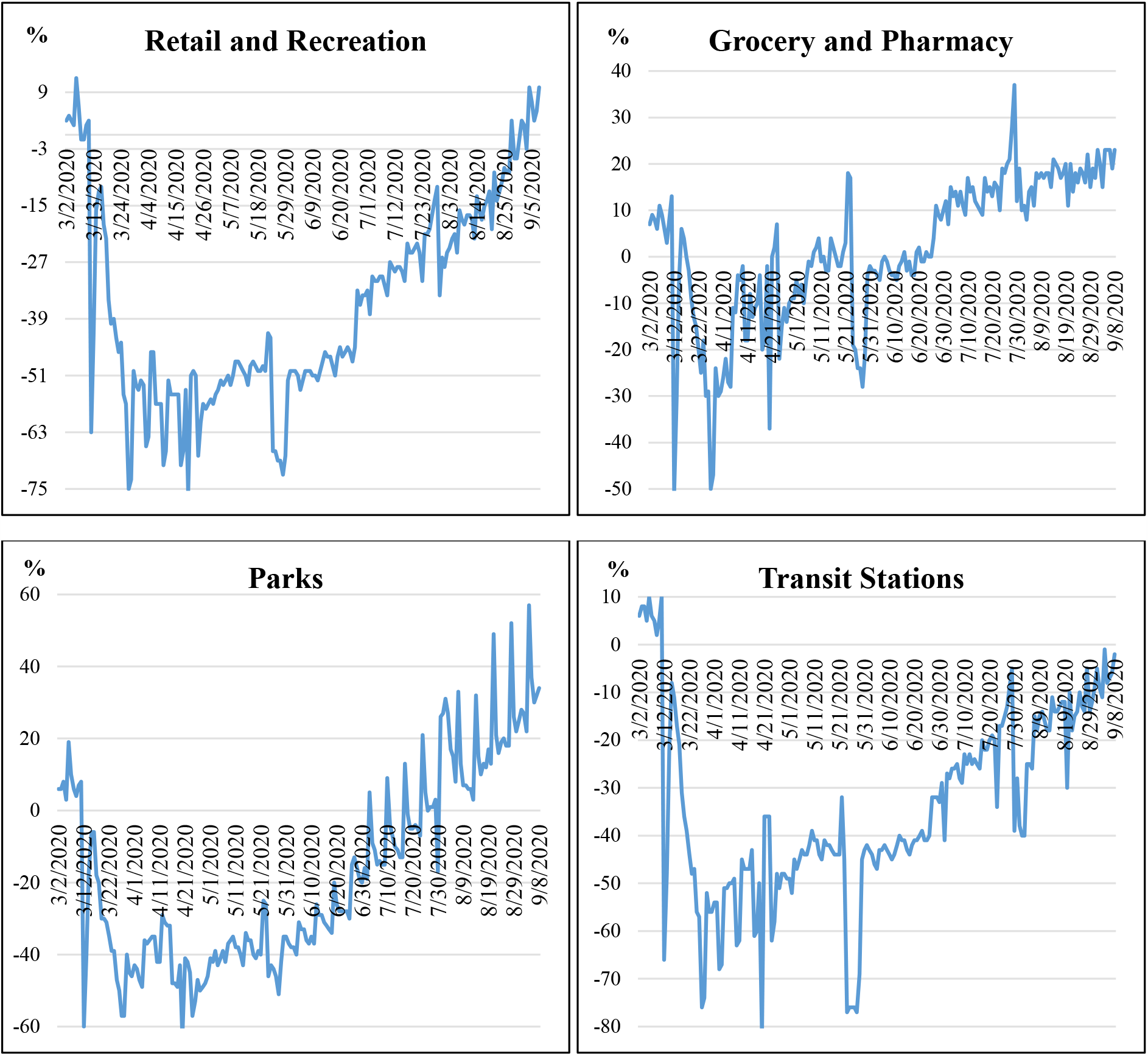
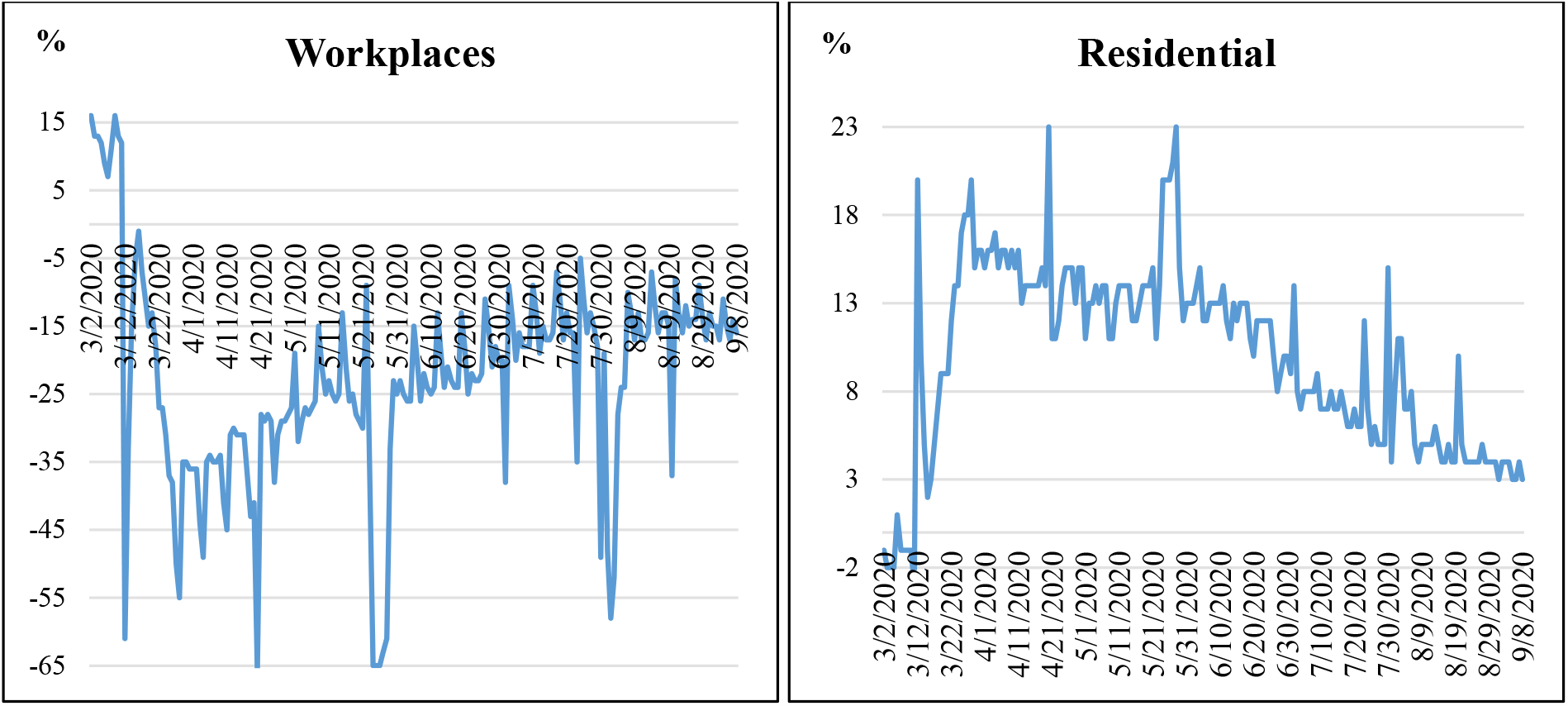
Mobility Trends’ Percarnt Change from Baseline

Clearly, the hike in the amount of individuals going out mirrors a spike in the amount of persons who have reportedly tested positive for the disease. Accordingly, the crowding that has occurred over the two weeks preceding the holy month of Ramadan has lead the curve of the infection to grow. It is worth mentioning that, right before the holy month, Egyptians were supposed to celebrate the Easter holiday on 20 April. However, amid fears of coronavirus outbreak, complete closure of public gardens and beaches was directed. Figure 3 shows clearly the huge recession in mobility on that day. Importantly, this day witnessed the biggest drop in the number of people going out. Visits to retail and recreation venues, parks, transit stations and workplaces were down 76%, 63%, 82% and 66% respectively. Afterwards, on 29 May 2020, the number of cases surpassed the 1,000 mark; this was due to the increased mobility of the Egyptians before and during Eid Al-Fitr.

As mentioned earlier, the highest number of infections taken place at the second half of June. Notably, ten days prior to this latter date, movement to retail and recreational outlets and parks was 2% and 6% higher on average than its counterpart was at the first weeks of June. At the same time, the number of people staying home has dropped. Foundationally, visits to grocery shops and pharmacies started to increase starting from the beginning of June marking a possible reason that individuals embarked on applying self-isolation at homes following the Egyptian Health Minister Treatment protocol.

Later and after about a three-month closure, by the end of June the government started reopening. After that, individuals started spending more time out and as a result time spent at home dropped by 5%. On the other hand, hanging to restaurants, cafes and other places increased 20% on average during July than June. Likewise, the portion of individuals visiting parks raised by 25% on average for the same period. At the same time, grocery stores and pharmacies witnessed 16% increase in visits. Additionally, use of transportation hubs jumped 18% on average. Despite resulting in upward movement as well as having Eid Al-Adha holiday, the number of infections kept on decreasing since Mid-July. This may be because an increasing number of Egyptians are taking social distancing measures seriously and committing to adhering to preventive measures to limit coronavirus outbreak as a result of increased level of awareness.

## 4. Methodology

Modeling and forecasting daily number of cases and deaths can help the health system provide services for the infected individuals and provide an insight for policy makers to design protective measures. Consequently, the econometric models could be of help in forecasting trajectories of the pandemic. In this study, both ARIMA model and ARDL model are employed to predict daily-confirmed cases and deaths of Covid-19.

### 4.1 Data

Speaking of the utilized data, a total number of 192 days (from March 2, 2020 to September 8, 2020) were used to develop the ARIMA and ARDL models of new cases and deaths. The Egyptian Ministry of Health and Population (MoHP) reports the official data regarding Covid-19 on a daily basis for instance, daily-confirmed cases, recoveries and deaths, as well as the total figures.

In 2020, Google released GCMR for many countries including Egypt. These reports aim to give vision about the changes in response to the different policies aimed at alleviating Covid-19 consequences. The reports provide mobility alternations over time across different sets of places such as; retail and recreation covering visits to restaurants, cafes, shopping centers, museums, libraries and movie theaters, groceries and pharmacies, parks, transit stations including subway stops and bus and train stations, workplaces, and residential places with respect to a baseline (Google LLC).

In other words, the dataset highlights the percent change in visits to different categories of places. It depicts how the frequency and/or length of visits to categorized places change compared to baseline days. The baseline day corresponds to the median value from the 5-week period Jan 3 – Feb 6, 2020 (Ibid). Using this data one can assess the effect of mobility patterns on the spread of Covid-19.

According to Lauer, et al. (2020), the median incubation duration is 5.1 days (95% Confidence Interval (CI), 4.5 to 5.8 days), and 97.5% of individuals who develop symptoms will do so within 11.5 days (CI, 8.2 to 15.6 days) of infection. As a result, it is to be expected that any changes in mobility will have a lagged effect on detecting the new infections. Consequently, lagged values of the mobility elements and indicators are estimated by means of time series models.

### 4.2 Models’ Estimation

Number of new cases (NCases) and new deaths (NDeaths) on daily basis are used in the estimation as dependent variables in two different models to be forecasted over two months (9 September - 7 November). Additionally, six independent variables – represent mobility alternations - are included in the econometric model of daily cases while one – new infections - is used for the deaths model.

#### 4.2.1 Estimation of Time Series Models

Many researchers have utilized ARIMA model to forecast the infection horizon of the virus in different countries (Dehesh, et al., 2020; Yousaf, et al., 2020; Kumar, et al., 2020; Benvenuto, et al., 2020; Saba & Elsheikh, 2020). Popularly known as the Box–Jenkins (BJ) methodology, the ARIMA methodology emphasizes on analyzing the probabilistic, or stochastic, properties of time series on their own under the philosophy *let the data speak for themselves* (Gujarati & Porter, 2009). Unlike the regression models, in which *Y*_*t*_ is explained by *k* regressors *X*_1_, *X*_2_, …., *X*_*k*_, the BJ-type time series models allow *Y*_*t*_ to be explained by past, or lagged, values of *Y* itself and stochastic error terms. The important point to note is that, the time series data used in ARIMA should have stationary and linear nature. Hence, a stationarity test is conducted: for instance, Autocorrelation Function (ACF), Correlogram, or the unit root test using the Augmented Dickey– Fuller (ADF) Test or the Phillips Perron Test.

The ARIMA (*p,d,q*) model includes an autoregressive (AR) model and a MA model where *p* denotes the number of autoregressive terms, *d* the number of times the series has to be differenced before it becomes stationary, and *q* the number of moving average terms. The chief tools in identifying the parameters of the model are the ACF, the Partial Autocorrelation function (PACF) and the resulting correlograms. Having chosen a particular ARIMA model, and having estimated its parameters, the next step is checking whether the chosen model fits the data reasonably well. All the models that passes the residual tests should be compared using Akaike information criterion (AIC). The model which has the least AIC is selected as the best model. Afterwards, the forecasting step based on the previously best-fit model is carried out.

#### 4.2.2 Estimation of Econometric Models

Until the beginning of the 1990s, static regression dominated the literature. It assumed that all the variables in the model are stationary. It has been proved that the macroeconomic data are mostly non-stationary; therefore, the OLS estimator does not produce reliable estimates and the regression tends to be spurious. Researchers use differenced variables in the model to obtain stationary variables, but in this case important information related to the long-run analysis is lost.

The co-integration approach is very attractive since it retains the long-run relations and obtains highly consistent parameters in the long run (Stock, 1987). However, there are integration and co-integration restrictions that the models have to overcome in order to apply this approach. Firstly, the ADF test (Dickey & Fuller, 1979) is used to examine the stationarity status of the variables in the different models. Pesaran, et al. (2001) proposed the ARDL approach of co-integration which can be applied and yields consistent estimates of the long-run parameters irrespective of whether the underlying variables are I(0), or I(1), or a combination of them. In addition, the ARDL approach permits different number of lags for each regressor to capture the data generation process in a general to specific framework (Feridun, 2009). The Monte Carlo results indicate that the ARDL approach works properly even when the model has endogenous regressors; the standard errors of the estimated coefficients are standard normal, therefore the standard critical values can be used and the diagnostic tests can be performed to evaluate the statistical performance of the models (Song, et al., 2009).

The ARDL approach involves several steps. First, the optimal number of lags for all level variables is selected, using the appropriate information criteria, mainly the AIC and Schwartz Information Criterion (SIC). The second step is the bounds test, which involves estimating the Conditional Unrestricted Error Correction Model (UECM) to test for the existence of a long-run steady state relationship between the dependent variable and all the explanatory variables. Then, we can proceed to estimate the UECM (p, q, m, n, s, v, u) as in Equation (1).

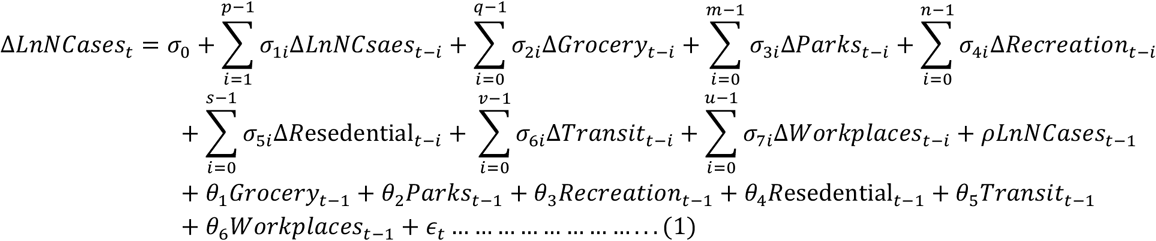

where p, q, m, n, s, v, u are the optimal lags of level of the regressors: LnNCases_t-i_, Grocery_t_, Parks_t_, Recreation_t_, Resedential_t_, Transit_t_, and Workplaces_t_ respectively, Δ is the first difference operator, and σ_0_ is a drift component. The left hand side of Equation (1) is the natural log of new cases (LnNCases_t_). The right hand side of the equation represents the explanatory variables in one lag in level, and in differences with the optimal lags for each variable; the independent variables represent six different measures of mobility alternations over time across different sets of places in Egypt. The parameters σ_si_ correspond to the short-run relations, whereas *ρ* and θ_*s*_ correspond to the long-run relations; ϵ_t_ is random errors.

The Wald or F-statistic is used to test the joint significance of lagged levels of the variables in the UECM, and determine the existence of the long-run equilibrium under the null hypothesis of no co-integration (H_0_: *ρ =* θ_1_ *=* θ_2_ *=* θ_3_ *=* θ_4_ *=* θ_5_ *=* θ_6_ *=* 0) against the alternative that a long-run relation exists (H_1:_ ρ ≠ θ_1_ ≠ θ_2_ ≠ θ_3_ ≠ θ_4_ ≠ θ_5_ ≠ θ_6_ ≠ 0) in Equation (1). However, as discussed by Pesaran, et al. (2001), both statistics have no standard distribution, irrespective of whether the regressors are purely I(0), purely I(1) or mutually co-integrated. Therefore, Pesaran, et al. (2001) computed two types of asymptotic critical values for a given significance level in the case of including and excluding trend. The first type assumes that all the variables are I(1), and the other assumes that all the variables are I(0). If the computed Wald or F-statistics exceed the upper critical value, the null hypothesis is rejected and the underlying variables are co-integrated. If the Wald or F-statistics are below the lower critical value, the null cannot be rejected and the variables are not co-integrated. Finally, if the Wald or F-statistic values lies between the two bounds, the test is inconclusive.

Once a long-run relationship has been established by the bounds test, the long-run relations can be estimated as 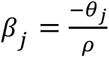. Since *j* represents the independent variables in Equation (1), *β*_*js*_ are the long-run coefficients of the different independent variables in the equation.

Finally, the models have to undergo several statistical checking such as autocorrelation, heteroscedasticity and stability test in order to ascertain their statistical reliability to be used in forecasting.

### 4.3 Forecasting Procedures

#### 4.3.1 Ex-Post Forecasting Procedures

An ex-post forecast (after the event forecast) is a forecast that is run in past periods for which actual values of the new cases and new deaths, in addition to the explanatory variables are available. The comparison of ex-post forecasts between different methods of estimation allows researchers to decide which method generates the best forecast, and therefore it can be used to produce ex-ante (before the event) forecasts (Nosier, 2012). Ex-post forecasting in the present study aims to evaluate the forecasting performance of the ARDL co-integration estimates and ARIMA (*p d q*) estimates for comparison.

The ARDL and ARIMA methods are re-estimated for the new cases and new deaths of Covid-19 models using daily data for the period from 2^nd^ of March to 25^th^ of August 2020. The estimated parameters are used to generate dynamic, ex-post forecasting over 14 days for the period (26^th^ of August – 8^th^ of September 2020) for the two estimated models. Dynamic forecasting calculates dynamic, multi-step forecasts starting from the first period in the forecast sample. In dynamic forecasting, previously forecasted values for the lagged dependent variables are used in generating forecasts of the current value. This choice will be available for our models, since the estimated equation contains dynamic components, such as lagged dependent variables and ARIMA terms.

The appropriate method of forecasting is chosen as the method, which has the least forecasting errors according to the forecasting error criteria, such as Root Mean Square Errors (RMSE), Mean Absolute Errors (MAE), Mean Absolute Percentage Errors (MAPE) and Symmetric Mean Absolute Percentage Errors (SMAPE).

Forecasting errors are defined as the differences between the actual and forecasted value over the period of the forecasting horizon, which are defined as:

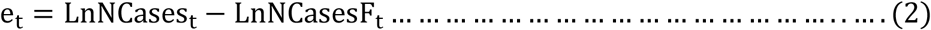

where LnNCases_t_ is the actual value of the natural log of new cases in time t, LnNCasesF_t_ is the forecasted value of the natural log of new cases in time t, *e*_*t*_ is the forecasting errors in time t. In theory, for a well specified model, it is expected that the forecasting error has a mean of zero over a certain forecasting horizon. However, very small forecasting errors can be obtained even if the models are not well specified as a result of the existence of positive and negative forecasting error values, which cancel each other out. To solve this problem, measures of forecasting errors accuracy have been improved and the errors of Equation (2) transformed either to squared values 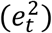 as in the RMSE in Equation (3), or to absolute values |*e*_*t*_| as in the MAE in Equation (4) (Song, et al., 2009).

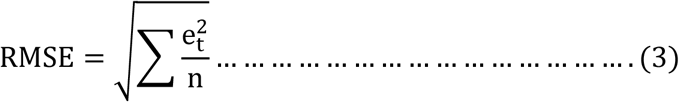

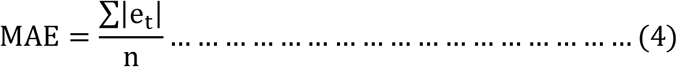

where n is the number of forecasts. The main difference between the two forecasting error measures is that the MAE gives equal weight to all errors, whereas the RMSE gives more weight to larger errors. Therefore, the RMSE is more sensitive to one extremely bad forecast (Li, et al., 2005). The MAPE is another error forecast measure in which the errors of the forecast are divided by the actual values of the dependent value, as in Equation (5), to produce unit independent measures (percentage errors), so one can compare the errors of fitted models that differ in level.

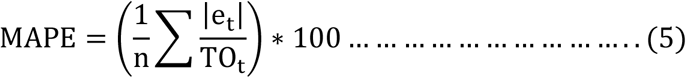

In the present study, the three measures are used to compare forecasting accuracy between different methods for the same model, whereas the MAPE is used to compare forecasting accuracy even across the two models. Moreover, Lewis (1982) suggested the following interpretation of the resulting MAPE values: Less than 10% is highly accurate forecasts, 10%-20% is a good forecasting, 20%-25% is a reasonable forecasting, 50% or more is an inaccurate forecasting (Lewis 1982, quoted in Tideswell, et al., 2001).

#### 4.3.2 Ex-Ante Forecasting Procedures

Ex-ante forecasting is very important for decision-making for both the private and public sectors. Generating accurate and valid forecasts requires well specified, theoretically consistent and statistically robust econometric methods (Nosier, 2012).

Statistical diagnostics and stability tests should be performed to examine the validity of the estimated ARIMA and ARDL in forecasting the future new cases and deaths of Covid-19 in Egypt. Moreover, ex-post forecasting comparison is implemented to select the preferred method for each model. The picked best-fit models are utilized to forecast daily cases and deaths for 60 days using the whole time span of 192 days. A longer forecast horizon would result in increasing errors of the forecast variables, resulting in lack of robustness and accuracy of the econometric forecast (Lee, 2005).

## 5. Results

In this section, the results of the estimated ARIMA and ARDL models are presented and discussed. Diagnostic tests are performed to the resulting estimates. If the used methods overcome the diagnostic and stability tests, we proceed by using the fitted models in generating ex-post and ex-ante forecasting of daily cases and deaths in Egypt over a period of two months from the 9^th^ of September to the 7^th^ of November as stated before.

### 5.1 Estimation Results

#### 5.1.1 Results of Time Series Models - ARIMA

ARIMA (4, 2, 1) is selected - according to the AIC- as the most appropriate specification to forecast the number of new cases of Covid-19 in Egypt, as illustrated in Figure 4 (A). The model as a whole is significant at the 1% level of significance and it has moderate explanatory power (R^2^= 0.613). The residuals of the model have no first order autocorrelation according to DW statistic (1.67), but the residuals are heteroscedastic (F-statistic of Autoregressive Conditional test of heteroscedasticity (ARCH) = 24.47, p-value = 0.000). The model is stable according to the Regression Equation Specification Error Test (RESET) (F-statistic = 0.551, p-value = 0.459). Therefore, except for heteroscedasticity, the ARIMA model has no econometric problems.

**Figure 4:**
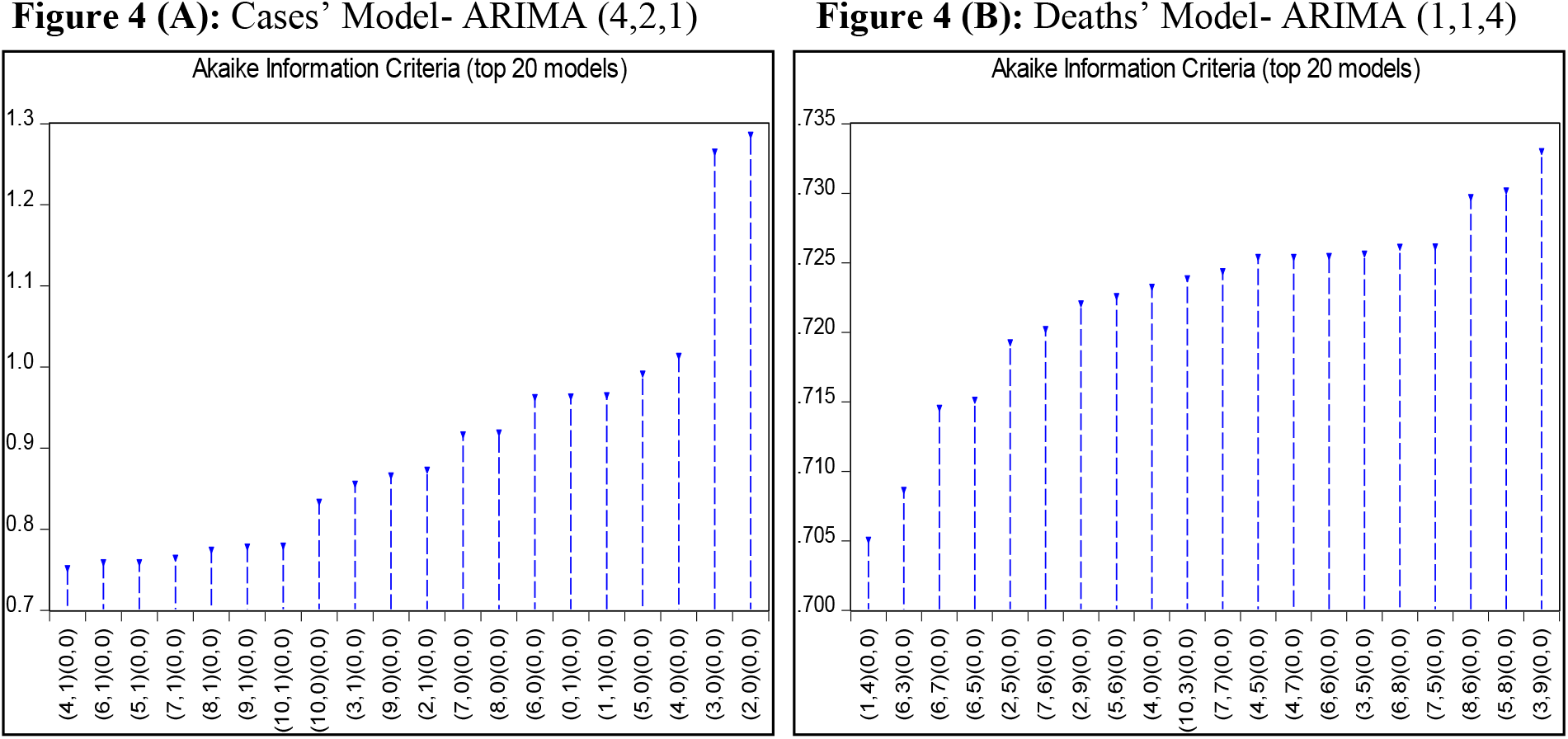
ARIMA Optimal Lags Selection Using AIC

In addition, ARIMA (1, 1, 4) is used to forecast the number of new deaths, as it is proved to have the best specification using the AIC, as shown by Figure 4 (B). The model is significant at the 1% level of significance, however, it has weak explanatory power (R^2^= 0.29). The residuals have no first order autocorrelation according to DW statistic (2.05). However, using ARCH test, the residuals are heteroscedastic (F-statistic = 8.514, p-value=0.004). Finally, the model is stable (F-statistic of RESET test=0.391, p-value=0.533). Therefore, the ARIMA model has no econometric problems, except for heteroscedasticity.

Solving for the problem of heteroscedasticity in the two models; ARIMA-ARCH model is utilized to obtain homoscedastic residuals. Both ARIMA and ARIMA-ARCH models are utilized in the ex-post forecasting to choose the highest forecasting accuracy models, which have the lowest forecasting errors.

#### 5.1.2 Results of the Econometric Models - ARDL

##### 5.1.2.1 Unit Root Results

The results of the ADF unit root tests are reported in Table 1. At the 5% level of significance, the results indicate that LnNCases_t_ is non-stationary, but stationary in first differences, *I*(1) variables, according to the appropriate trend-specification. All the other variables in the two models are found to be stationary variables, *I*(0). However, we proceed by estimating the co-integration relation of the two models, since we use the ARDL technique which permits a co-integration relationship to exist irrespective of whether variables have the same integrating order or not.

**Table 1:** Unit Root Tests According to the Appropriate Deterministic Trend

| Variables | Trend Specification | ADF | Variables | Trend Specification | ADF |
| --- | --- | --- | --- | --- | --- |
| $\text{LnNCases}_t$ | Constant +Trend | -1.206<br>(0.906) | $\Delta \text{LnNCases}_t$ | None | -3.637<br>(0.000) |
| $\text{LnNDeath}_t$ | Constant | -3.305<br>(0.016) | $\text{Residential}_t$ | Constant + trend | -3.797<br>(0.019) |
| $\text{Grocery}_t$ | Constant +Trend | -6.579<br>(0.000) | $\text{Transit}_t$ | Constant + trend | -4.564<br>(0.002) |
| $\text{Parks}_t$ | Constant+ Trend | -4.146<br>(0.007) | $\text{Workplaces}_t$ | Constant | -5.717<br>(0.000) |
| $\text{Recreation}_t$ | Constant + Trend | -4.290<br>(0.004) | | | |

##### 5.1.2.2 Bounds Test Results

The first step in ARDL approach of co-integration is to determine the optimal lags of the variables included in the models. Ten lags are set as a maximum lag length, and different ARDL specifications are chosen separately for each model and each variable according to the AIC as illustrated in Figure 5. Therefore; ARDL (10, 3, 0, 3, 2, 4, 1) has been selected as the best-fit model to forecast daily new cases, while, ARDL (9, 0) has been preferred for predicting daily new deaths.

**Figure 5:**
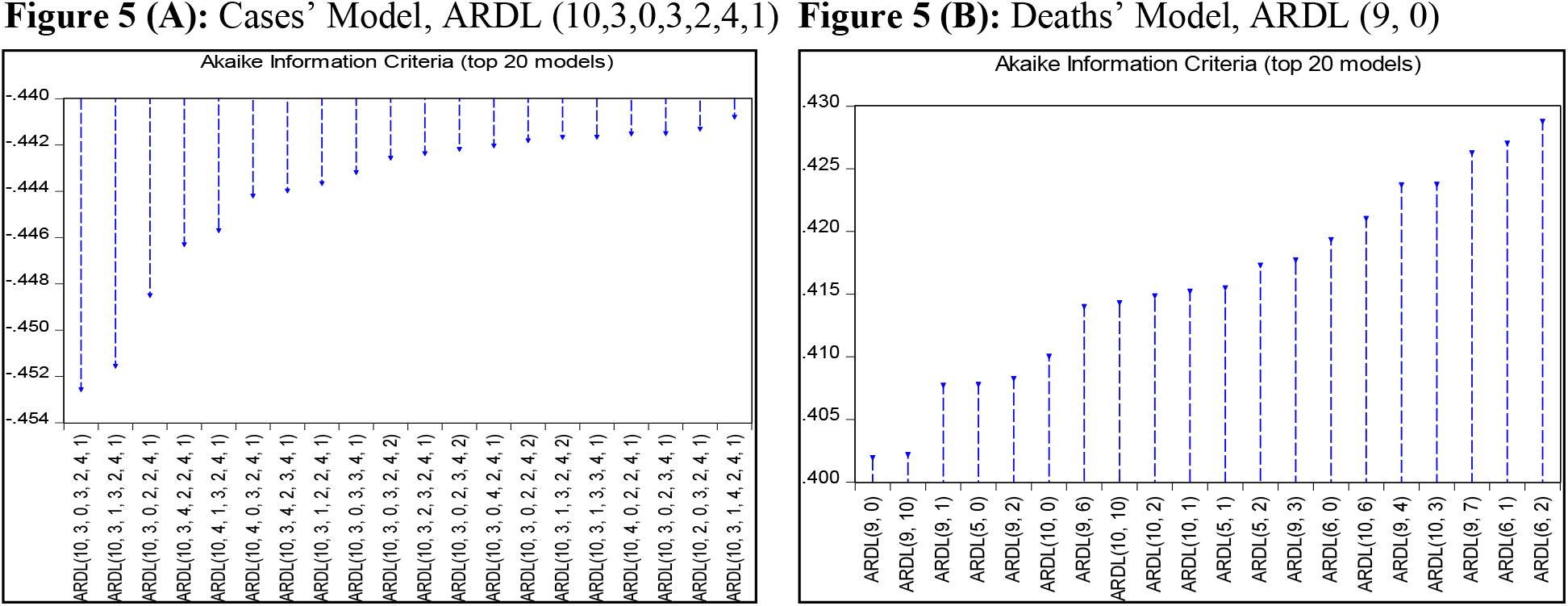
ARDL Optimal Lags Selection Using AIC

The second step of ARDL approach is to test the existence of the long-run Equilibrium relationship–co-integration relationship– between LnNCases and its determinants in the first model, and between LnNDeaths and its determinants in the second model using the bound test. The results of bounds tests (computed F-statistics) for the two models are presented in Table 2.

**Table 2:** The Results of F-Statistics for Co-Integration Relationship

|  | Cases' Model |  |  | Deaths' Model |  |  |
| --- | --- | --- | --- | --- | --- | --- |
| Critical value | Significance level |  | F-Statistic | Significance level |  | F-Statistic |
|  | 1% | 5% |  | 1% | 5% |  |
| Minimum value<br>I (0) | 3.15 | 2.45 | 7.848 | 6.84 | 4.94 | 13.648 |
| Maximum value<br>I (1) | 4.43 | 3.61 |  | 7.84 | 5.73 |  |

Note: The critical value of F is calculated at 6 independent variables in the Cases' model, and one independent variable in the Deaths' model. Constant without time trend is included in both models.

The null hypothesis of no co-integration is rejected in the two models, indicating that there is a co-integration relation at the 1% level of significance for the two models over the period of the study. Therefore, we can proceed by estimating the long-run relationships between these variables.

##### 5.1.2.3 Estimating the Long-Run Equilibrium Relationships

Having determined the best ARDL specification for the two models and the long-run co-integration relation is detected, long-run steady-state parameters were estimated and examined statistically in the two models as illustrated in Table 3.

**Table 3:** Long-Run Results of ARDL Co-Integration Approach

| Independent Variables | Cases' Model |  | Independent Variable | Deaths' Model |  |
| --- | --- | --- | --- | --- | --- |
|  | Coefficient | T-statistics |  | Coefficient | T-statistics |
| Grocery | 0.137<br>(0.000) | 3.857 | LnNCases | 0.716<br>(0.000) | 6.417 |
| Parks | -0.078<br>(0.013) | -2.520 |  |  |  |
| Recreation | 0.296<br>(0.001) | 3.346 |  |  |  |
| Residential | 0.089<br>(0.702) | 0.383 |  |  |  |
| Transit | -0.709<br>(0.000) | -3.630 |  |  |  |
| Workplaces | 0.211<br>(0.000) | 4.333 |  |  |  |
| Goodness of fit & diagnostic tests |  |  |  |  |  |
| R<sup>2</sup> | 0.981 |  | R<sup>2</sup> | 0.891 |  |
| Adj. R<sup>2</sup> | 0.977 |  | Adj. R<sup>2</sup> | 0.884 |  |
| Durbin-Watson stat | 2.026 |  | Durbin-Watson stat | 2.008 |  |
| B-G LM (F-statistics) | 0.457<br>(0.500) |  | B-G LM (F-statistics) | 0.113<br>(0.894) |  |
| ARCH hetero (F-statistics) | 0.627<br>(0.429) |  | ARCH hetero (F-statistics) | 8.103<br>(0.005) |  |
| RESET (F-statistics) | 3.200<br>(0.076) |  | RESET (F-statistics) | 2.557<br>(0.081) |  |

Note: B-G LM test is the Breusch-Godfrey Lagrange Multiplier statistics for residual autocorrelation. P-value between Brackets

The models perform reasonably. The adjusted R^2^ is very strong in both models, indicating strong explanatory power and the diagnostic tests indicate no statistical problems, except for heteroscedasticity in the new deaths’ model. White-Hinkley (HC1) heteroscedasticity consistent standard errors and covariance is used to control the problem of heteroscedasticity in this model. Moreover, the two models are stable. Therefore, forecasting using these models can be performed accurately.

For the cases’ model, all the long-run coefficients are significant at the 1% level of significance - but the residential variable is not significant - indicating that mobility of population is affecting the incidence of new cases of Covid-19 significantly over the period of the study. As expected, a 1% increase in visits to grocery and pharmacy places, recreation areas, and workplaces compared with the base line is associated with higher incidence of Covid-19 by 14%, 30%, and 21% respectively at the 1% level of significance. However, the long-run coefficients of parks and transit have an unexpected negative sign. In the deaths’ model, an increase in new cases by 1% tends to increase the new deaths by 0.72% on average in the long run.

Therefore, the spread of Covid-19 is caused by increasing mobility, especially to recreation areas-such as restaurants, cafes, shopping centers, theme parks, museums, libraries and movie theaters- and workplaces. Moreover, the spread of Covid-19 is more likely to increase the possibility of deaths.

### 5.2 Forecasting Results

#### 5.2.1 Ex-Post Forecasting Results

Speaking of the daily new cases, as illustrated before, ARIMA (4, 2, 1) was chosen as the best-fit model among other specifications. Regarding ARDL models, based on the same criterion, Dynamic ARDL (10, 3, 0, 3, 2, 4, 1) was selected as the best model. Concerning the daily deaths, the selected model is ARIMA (1, 1, 4) while Dynamic ARDL (9, 0) was picked. ARIMA-ARCH method for new cases and deaths models is also estimated and evaluated to treat the problem of heteroscedasticity in the ARIMA models.

The performance of the chosen models of daily cases and deaths respectively are compared using different measures of forecasting error accuracy. Table 4 shows the forecast error of these models in terms of RMSE, MAE, MAPE and SMAPE over the period of 14 days.

**Table 4:** Forecasting Error Criteria of all Models (26 August - 8 September)

|  | Cases' Model |  |  |  | Deaths' Model |  |  |  |
| --- | --- | --- | --- | --- | --- | --- | --- | --- |
| Forecasting Model | RMSE | MAE | MAPE | SMAPE | RMSE | MAE | MAPE | SMAPE |
| **ARIMA** | 0.487 | 0.446 | 8.447 | 8.891 | 0.443 | 0.409 | 13.736 | 14.876 |
| **ARIMA-ARCH** | 0.869 | 0.842 | 16.188 | 17.732 | 0.228 | 0.196 | 6.730 | 6.869 |
| **Dynamic ARDL** | 0.694 | 0.675 | 12.929 | 13.876 | 0.230 | 0.181 | 6.287 | 6.336 |

It is noticed that based on the different error measures, the ARIMA model is the best to forecast daily cases, whereas Dynamic ARDL is the most accurate one for daily deaths. Moreover, the MAPE indicates that only ARIMA method has highly accurate forecasts in the new cases model, with error magnitude less than 10%, whereas both ARIMA-ARCH and Dynamic-ARDL obtained a good forecast according to Lewis (1982) criterion. For the death model, the opposite is true since Dynamic ARDL and ARIMA-ARCH have highly accurate forecasts, whereas ARIMA only has a good forecast. Comparison among models can be also done across models using the MAPE since it is unit independent measures. Dynamic ARDL of the new deaths is the best one with the least forecasting errors.

The graphical investigation confirms the latter results. Figure 6 and Figure 7 compare the results of the preceding methods for the ex-post period (26 August - 8 September), and illustrate that ARIMA model provides the closest results to the actual data for daily cases, whereas Dynamic ARDL is the closest to the actual daily deaths.

**Figure 6:**
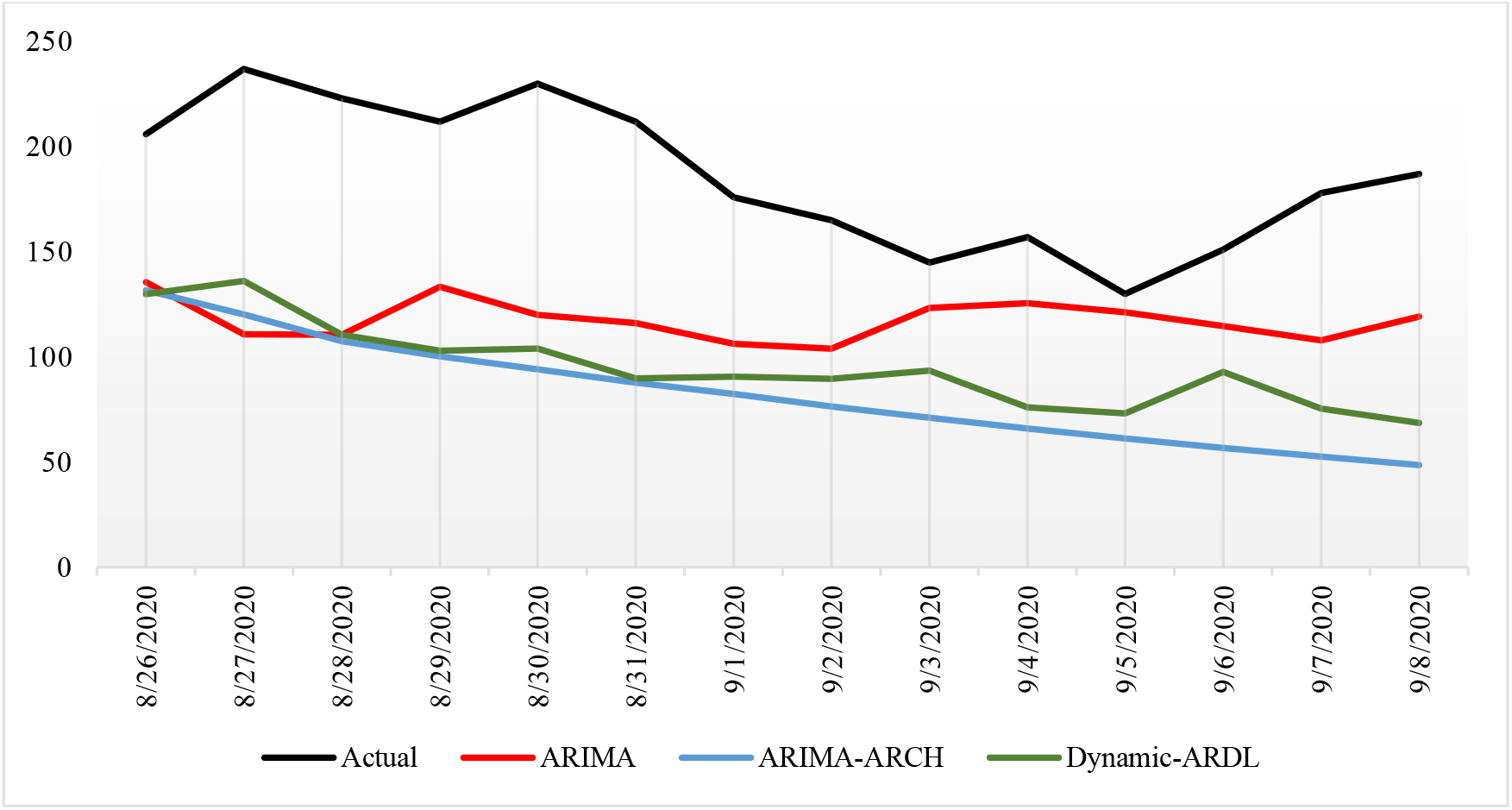
Ex-Post Forecasting of Daily Cases Using Different Methods

**Figure 7:**
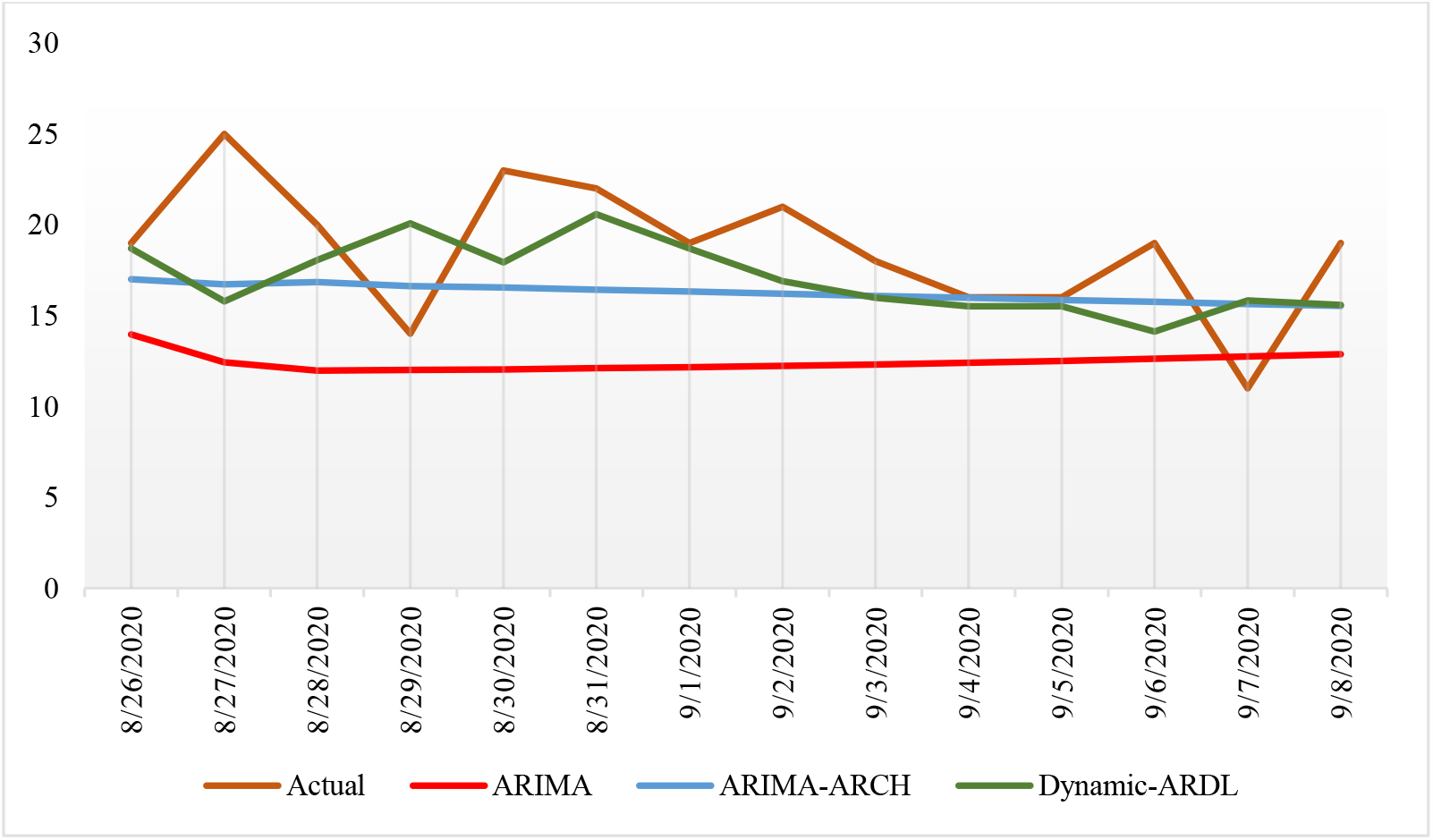
Ex-Post Forecasting of Daily Deaths Using Different Methods

#### 5.2.2 Ex-Ante Forecasting Results

To generate ex-ante forecasting using econometric methods, the future values of the explanatory variables have to be forecasted first. The ARIMA method is used to forecast the future values of the explanatory variables of the two models before proceeding to forecasting the values of new cases and deaths. Table 5 illustrates the predicted values of infections and deaths at the end of each month of the forecasting horizon using different forecasting methods for each model for comparison.

**Table 5:**
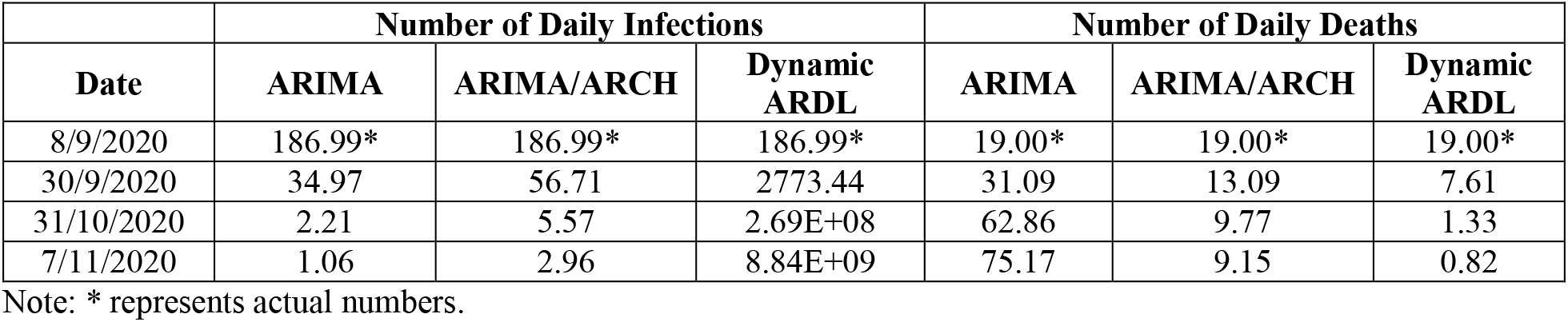
Ex-Ante Forecasts of Infections and Deaths (9 September-7 November)

| Date | Number of Daily Infections |  |  | Number of Daily Deaths |  |  |
| --- | --- | --- | --- | --- | --- | --- |
|  | ARIMA | ARIMA/ARCH | Dynamic ARDL | ARIMA | ARIMA/ARCH | Dynamic ARDL |
| 8/9/2020 | 186.99* | 186.99* | 186.99* | 19.00* | 19.00* | 19.00* |
| 30/9/2020 | 34.97 | 56.71 | 2773.44 | 31.09 | 13.09 | 7.61 |
| 31/10/2020 | 2.21 | 5.57 | 2.69E+08 | 62.86 | 9.77 | 1.33 |
| 7/11/2020 | 1.06 | 2.96 | 8.84E+09 | 75.17 | 9.15 | 0.82 |

Note: \* represents actual numbers.

First, the outcomes of the selected methods of forecasting for the anticipated daily cases for 60 days (9 September-7 November) are depicted in Figure 8.

**Figure 8:**
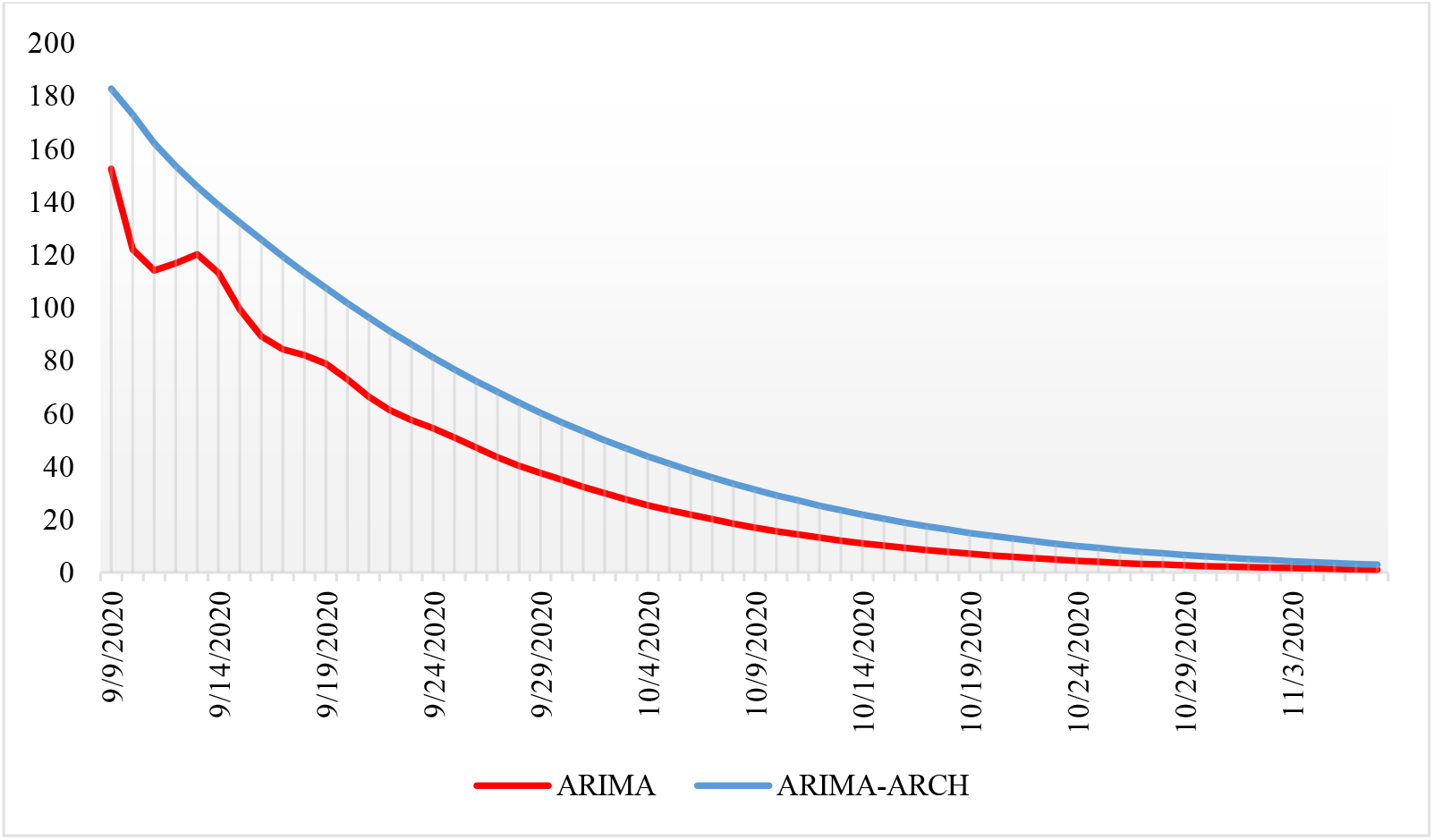
Forecasted Daily Cases Using Different methods (9 September-7 November)

Although the ARIMA model has the highest forecast accuracy for the new cases model, we display it along with ARIMA-ARCH only for comparison purposes. As demonstrated by the hereinabove Figure, the two-time series methods anticipate a decrease in the daily infections although ARIMA-ARCH model predicts less fall than ARIMA. The first anticipates new infections to drop from 187 cases on September 8, 2020 to reach 3 cases by November 7 while the latter forecasts only 1 new case by that day. To add more, total number of infections on 7 November 2020 are anticipated to reach 102,352 and 103,365 cases by ARIMA and ARIMA-ARCH respectively. That is, a 2.12% and 3.13% increase in the number of infections is predicted.

According to Dynamic ARDL, the daily cases soar continually up to the end of the forecasting period to reach millions of cases. Caution needed when interpreting the latter results since the future values of the mobility variables are estimated without any restrictions on the full capacity of the different mobility places over time. Therefore, the increasing rate of movements to these places increases the number of new cases strongly as well.

In the same way, Figure 9 presents the results of the fitted models for daily kills. The results of dynamic ARDL (the best forecasting model) and ARIMA-ARCH revealed that daily deaths tend to decline over time across the period of forecasting.

**Figure 9:**
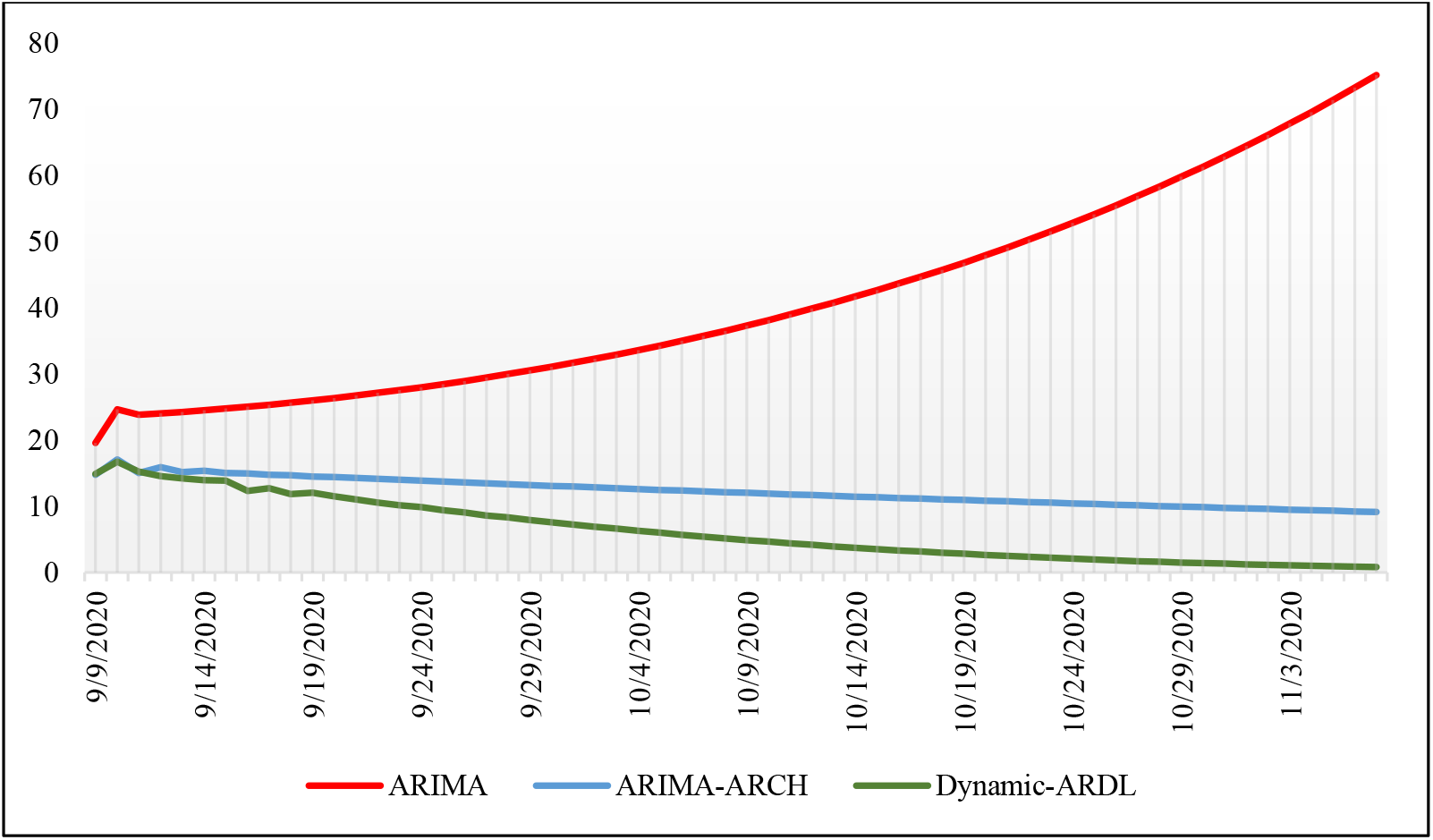
Forecasted Daily Deaths Using Different Methods (9 September-7 November)

Despite anticipating a decreasing mortality rate, there is a small difference between the two models’ outcomes. For instance, it is predicted that the number of deaths tends to fall seriously from 19 case at September 8 to 1 and 9 deaths at the end of the forecasting period (7^th^ November) according to the dynamic ARDL and ARIMA-ARCH models respectively. In sum, total death toll is predicted to reach 5,938 (Dynamic ARDL) or 6,294 (ARIMA-ARCH) at the end of the forecasting horizon. To put it differently, the number of deaths is foreseen to rise by 13.21% or 6.80%. On the other side, mortality rate related to Covid-19 is expected to increase continuously using ARIMA forecasting model to reach 75 daily deaths, and 8,024 as total deaths by the end of the forecasting period.

## 6. Discussion

Although more data and information are needed, econometric models can be useful in predicting confirmed infections and deaths as well as future pandemic spread rates if the spread of the virus does not change very oddly. Undoubtedly, it is globally well known that this virus is new and has the potential to be highly transmissible in addition to growing fears of a powerful second wave. This ability may influence all expectations, but to our knowledge and time of estimation these models are among the best and most recent.

Regarding the current situation in Egypt, the last month has witnessed the rise of the infection rate by 0.29% whilst the death rate has fallen by 0.27% and the recovery rate has increased by 23.54%. Obviously, this is due to the serious early list of closures and policies imposed by the Egyptian government which aimed at encouraging citizens to stay at home to stop the spread of infections.

Additionally, three-month after directing the previously mentioned closure at end of June the government started reopening to avoid GDP decline, income losses and poverty increase. Despite resulting in upward movement, the number of infections kept on decreasing since mid-July. This may be because an increasing number of Egyptians are taking social distancing measures seriously and committing to preventive measures to limit coronavirus outbreak as a result of increased level of awareness.

Chiefly, the results of the utilized models conclude that mobility of population is affecting the incidence of new cases of Covid-19 significantly. As expected, an increase in mobility related to grocery and pharmacy places, recreational outlets, and workplaces by 1% is associated with higher incidence of Covid-19 by 14%, 30%, and 21% respectively. Therefore, the spread of Covid-19 is caused by increasing mobility, especially recreation places such as restaurants, cafes, shopping centers, theme parks, museums, libraries and movie theaters, in addition to workplaces. Moreover, the spread of Covid-19 by 1% more tends to increase the possibility of deaths by 0.72% on average in the long run.

Ex-post forecasting is performed for a period of two weeks (26 August - 8 September) using both time series and econometric techniques to choose the most accurate forecast for each model to be used in the ex-ante forecasting of the new cases and new deaths models. ARIMA is the best for the new cases model, while Dynamic ARDL is the best for the new deaths model. Results of other forecasts, which obtained using different techniques, are also shown for comparison.

The ex-ante forecasting results using different approaches reveal two different suggested Scenarios to the future Covid-19 epidemic. Table 6 illustrates the growth rates of the forecasts of infections and deaths at each month of the forecasting horizon using the two Scenarios, along with the actual growth rates. First and the more accurate Scenario - according to our forecasting accuracy measures - is the Optimistic Scenario. It is expected that the number of infections by the virus is decreasing continuously to reach only one case daily by total number of 102,352 cases at the 7^th^ November. Moreover, the anticipated number of kills due to Covid-19 is predicted to fall seriously from 19 case at September 8 to 9 deaths by total of 5,938 deaths at the end of the forecasting horizon. Thus, the results show possible containment of the pandemic. Figure 10 illustrates this optimistic Scenario along with the actual data of the new infections and deaths of Covid-19 over the period (15^th^ of March – 7^th^ of November).

**Table 6:** Growth Rates of Number of Infections and Deaths (Actual and Forecasts)

| Date | Optimistic Scenario |  | Pessimistic Scenario |  |
| --- | --- | --- | --- | --- |
|  | Daily Growth Rate of Infections (%) | Daily Growth Rate of Deaths (%) | Daily Growth Rate of Infections (%) | Daily Growth Rate of Deaths (%) |
| 15/3 - 30/6 | 4.3* | 4.2* | 4.3* | 4.2* |
| 1/7 - 8/9 | -3.0* | -2.1* | -3.0* | -2.1* |
| 9/9 - 30/9 | -6.5 | -3.0 | 13.4 | 2.1 |
| 1/10 - 31/10 | -8.4 | -5.3 | 43.4 | 2.2 |
| 1/11 - 7/11 | -8.6 | -5.7 | 54.3 | 2.2 |

Note: \* represents growth rate of actual numbers.

**Figure 10:**
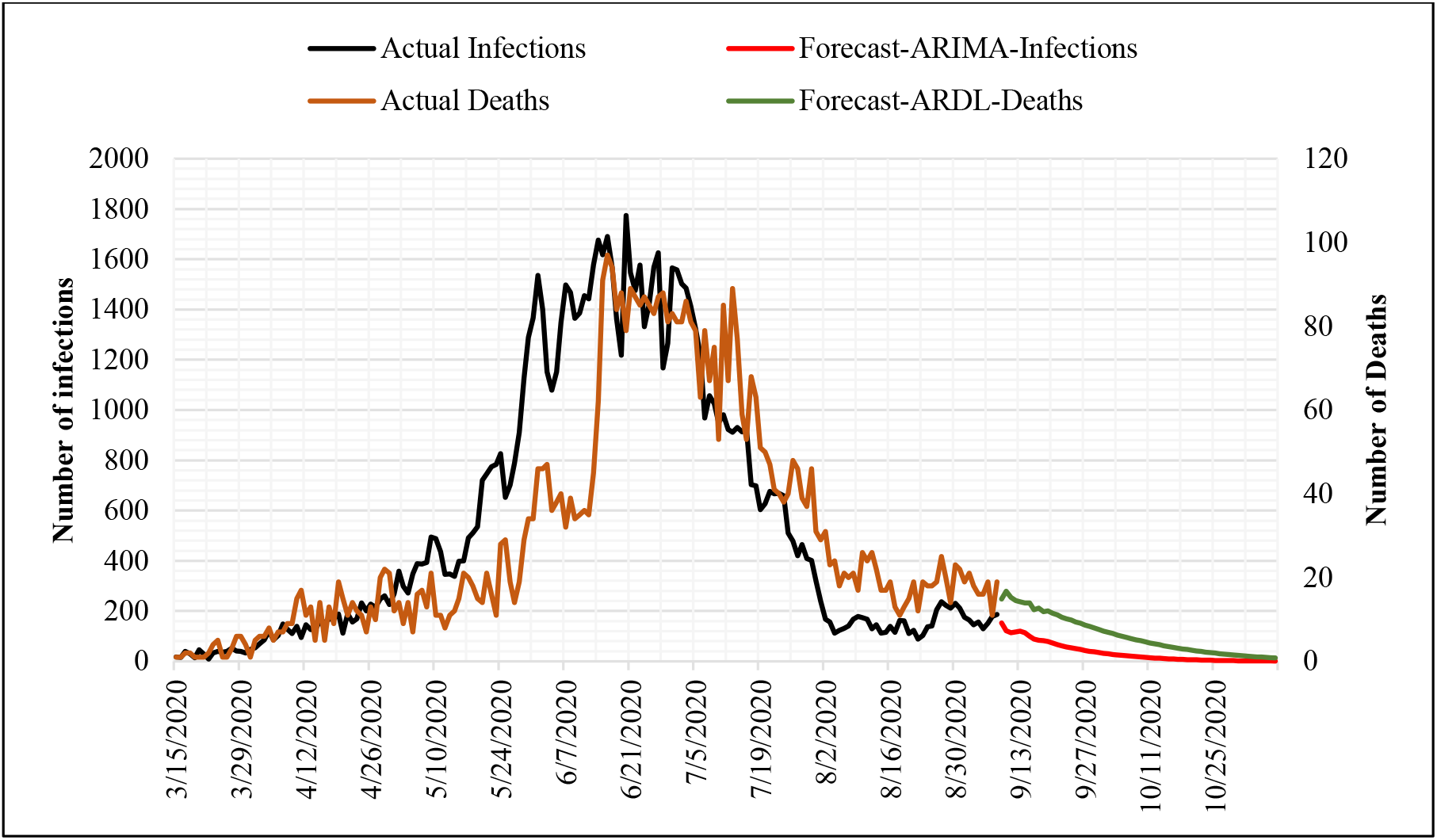
The Actual and Forecasts of New Infections and Deaths of Covid-19 “The optimistic Scenario”

On the other side, other forecasting techniques - although less accurate ones – reveal a sharp increase in infections by the coronavirus to reach millions of cases on the 7^th^ of November. This pessimistic Scenario suggests that the majority of persons in Egypt is expected to be infected by the virus at the end of the forecasting period (Leading to herd immunity). However, this huge increase in the number of infections, 33.6% daily growth rate on average over the forecasting period, is associated with much less growth rate of the number of deaths over the same forecasting horizon. The latter is only 2.3% daily on average of all the forecasting period to reach its maximum of 75 daily deaths by total of 8,023 deaths at the end of the forecasting horizon. Moreover, the growth rate of deaths is increasing steadily across time, while the daily growth rate of infection is increasing sharply from month to month as illustrated in Table 5. The latter result suggests huge spread of the coronavirus with less risk of deaths in the coming two months. So, quicker spread, with less number of killed persons.

Even if the pessimistic Scenario is less likely to occur according to our forecasting accuracy measures, policy makers have to put it into consideration for best control of the epidemic.

Since the future of Covid-19 depends on the decisions made in addition to community engagement practices among others, countries should plan and be ready for any scenario. To maintain the expected trajectory of the pandemic in Egypt, many restrictions must be continued and emergency mechanisms need to be considered. Regarding the community role, social distancing measures must be strictly adhered to. In some countries such as England, Hong Kong, Singapore, and Norway, a 1-meter distance is recommended while, Germany and Spain endorsed a 1.5 meter and a 2-meter space is advised in Japan and South Korea (Han, et al., 2020). Besides, following the declared health precautions and rules to ensure that infection is avoided or transmitted is substantial. Consequently, maintaining good hygiene is of great importance, that is, wearing masks everywhere, frequently washing hands and staying home in case of being sick. It should also be noted that it is possible that this expected pattern will change, but this depends on the actions of the people that facilitate the work of the health system in each country.

Speaking of the role of the government, plans of re-imposing lockdown are recommended, upon setting certain epidemiological threshold. To put it another way, some governments have marked 50 new cases per 100,000 residents per state for 7 continuous days as the threshold for re-closing. It is also recommended continuing working from home for those who are more vulnerable to infection while promoting distancing and hygiene rules for workers who will be physically present at workplaces. For the purpose of preventing more infections to happen, it is recommended to enhance the effective find, test, trace, isolate, and support systems. Hence, temperature checking needs to be generalized before entering different places in addition to using the sterilization gates.

As a trial to prevent predicted deaths or to deal with possible second wave of infections especially following reopening, a support to the Egyptian health-system is crucial. That is increasing the number of hospitals equipped with intensive care units and ventilators for patients. Additionally, providing protective equipment for the medical sector staff and health-care workers.

Moreover, it is advised to continue restricting flights from severely infected countries. While screening individuals coming from other countries and quarantine measures are essential to pick potential infections and prevent more transmission. Regarding internal mobility, additional trains and buses can be recommended to run to alleviate overcrowding, especially at peak times. Equally important is keeping restrictions on all different social gatherings to limit the further spread of the virus.

These recommended strategies should support Egypt in stopping viral spread of the virus and avoid returning to full lockdown.

## Data Availability

the data is available upon request

## Notes

### Competing Interest Statement

The authors have declared no competing interest.

### Funding Statement

no funding

